# SIGNS OF CENTRAL SENSITIZATION IN PATIENTS WITH MUSCULOSKELETAL DISORDERS AND CHRONIC COMORBIDITIES: A SYSTEMATIC REVIEW AND META-ANALYSES OF OBSERVATIONAL STUDIES

**DOI:** 10.1101/2023.02.02.23285365

**Authors:** Mattia Sgarbi, Davide D’Alessandro, Matteo Castaldo, Daniel Feller

## Abstract

**Objectives:** this review aimed to investigate the presence of signs of central sensitization in patients with musculoskeletal disorders and associated chronic comorbidities.

**Methods:** we conducted a systematic review (prespecified protocol CRD42021228970). Two authors independently searched for primary studies published between 2000 and 2021 in Web of Science and PubMed databases. We searched for studies that investigate the presence of signs of central sensitization in patients with musculoskeletal disorder or migraine and a chronic comorbidity. Two authors independently evaluated the methodological quality of the included studies using the “The Joanna Briggs Institute Critical Appraisal tools”. When studies were judged homogenous enough, we performed a random effect meta–analysis.

**Results:** we included 14 observational studies. Overall, patients with musculoskeletal disorders or migraine with an associated comorbidity showed more signs of central sensitization compared with healthy subjects.

The quantitative analysis showed that patients with temporomandibular disorders and chronic comorbidities, compared to patients with temporomandibular disorders without comorbidites, have a decreased pressure pain thresholds measured in the masseter area [SMD: -0.52; CI 95%: - 1.02 to -0.03; I^2^: 67%] and in the trapezius area [SMD: -0.55; CI 95%: -0.96 to -0.14; I^2^: 0%].

Patients with migraine, chronic low back pain or rheumatoid arthritis and associated fibromyalgia present more signs of central sensitization, measured in different modalities, than subjects without comorbidity.

**Conclusions:** we demonstrated that, in general, patients with musculoskeletal disorders and an associated comorbidity showed an increased incidence of signs of central sensitization compared to healthy subjects and patients with musculoskeletal disorders without comorbities.

## 1. Introduction

Musculoskeletal disorders (MSK-D) (e.g., osteoarthritis (OA), low back pain (LBP) and rheumatoid arthritis (RA)) are among the leading causes of long term pain and physical disability in the general population.^1,2^

In MSK-D, signs of central sensitization (CS) are often found,^3–6^ contributing to the high rate of physical disability associated with these pathologies. CS is defined by the International Association for the Study of Pain (IASP) as an *“increased responsiveness of nociceptive neurons in the central nervous system to their normal or subthreshold afferent input”*.^7^ Signs of CS are also found in subjects that have experienced acute injuries such as trauma,^8^ surgery^9,10^ or sprains,^11^ and it is believed to be one of the main causes of the persistence of long lasting symptoms in MSK-D^12–17^ and in chronic systemic inflammatory diseases.^3^

As reported by Curatolo and Arendt-Nielsen,^18^ currently, there is not a gold standard measurement method for assessing CS, and the current knowledge on the presence and mechanisms of central hypersensitivity stems mostly from basic and laboratory research. Humans’ very limited accessibility of nociceptive pathways is an obvious constraint in the clinical research.^18^ Therefore, the main evidence for central hypersensitivity in human pain is deduced from indirect measures:^18^ typically, signs and symptoms of CS include hyperalgesia, allodynia, widespread pain, and other such as fatigue, sleep disturbances, headache and intestinal problems.^19,20^

In clinical practice, the most valuable tools used to identify patients with signs of CS are (as summarized by Den Boer et al.^21,22^): the central sensitization inventory (CSI), the pressure pain thresholds (PPT) (where an algometer performs pressure stimulation on different parts of the body, and PPT are measured), and monofilaments (where a thick thread applied several times with a time interval is used to measure temporal summation (TS)).

MSK-D, such as LBP, OA, and RA, often coexist with chronic medical conditions (i.e. comorbidities), such as hypertension, type 2 diabetes, depression, headache, overweight and obesity.^2,23,24^ The general health state, and consequently the presence chronic comorbidities, have emerged as two important factors in maintaining CS.^25^ Hence, patients with MSK-D and associated comorbidities could be at an increased risk of developing hypersensitivity to pain compared to patients with only a MSK-D or healthy subjects, as stated by the review of Costa et al.^26^ on temporomandibular disorders (TMD). However, in the literature there are not data concerning the degree of CS in other types of MSK-D with chronic comorbidities. It could be important to understand if this characteristic is common to all patients with MSK-D and associated comorbidities, because CS could be one of the reasons for the maintenance of long-term musculoskeletal pain and symptoms in patients with comorbidities.

Therefore, this systematic review (SR) aimed to investigate and summarize the literature concerning the presence of signs of CS in patients with MSK-D associated with chronic comorbidities.

## 2. Materials and methods

### 2.1 Study design and protocol

We registered the present SR protocol in PROSPERO (CRD42021228970). We reported the following manuscript according to the “PRISMA 2020 statement”.^27^

### 2.2 Eligibility criteria

We included observational studies (i.e., cross-sectional studies, case-control studies, cohort studies) and clinical trials that measured signs of CS in patients at least 18 years old with a MSK-D (either acute or chronic) associated with one or more chronic comorbidities. We considered two type of headache, migraine and tension type headache (TTH), in the group of MSK-D because, despite being a neurological disorders, often have associated muscoloskeletal dysfunctions.^28–30^ In addition, physiotherapy is increasingly used as an effective alternative to drugs to manage pain and improve disability in these types of headache.^31,32^ As comorbidity, we mean any type of chronic pathology, the pathology can involve any system. For this reason, the same chronic MSK-D (better known as persistent MSK-D) (e.g. OA, RA, chronic LBP (CLBP)) and any type of headache were considered to be compatible also as associated comorbidity in this SR. Fibromyalgia (FMS), as chronic fatigue syndrome and irritable bowel syndrome, was not considered a MSK-D but was considered comorbidities because these three conditions are part of the central sensitivity syndrome (CSS).^33^ This term describes a group of medically indistinct (or nonspecific) disorders, for which CS may be a common part of their etiology.^33–35^ Furthermore, we considered insomnia, sleep disorders, psychological trauma, and drug abuse as comorbidity only if a medical doctor diagnosed them according to the “Diagnostic and Statistical Manual of mental disorders-5”^36^ and therefore it was not enough to be self-reported by the patients. The diagnosis of the comorbid chronic medical condition had to be done before assessing the signs of CS. To be included, studies had to compare patient samples with a group composed of healthy subjects, patients with only MSK-D, or patients with a general chronic comorbidity.

We considered signs of CS the ones measured only with the methods indicated by den Boer et al.^21^

### 2.3 Search strategy and study selection process

We searched MEDLINE (through PubMed) and WEB OF SCIENCE (WoS), from 2000 to December 2021 since the reporting of CS in MSK-D has changed and increased substantially within the last 20 years. Studies must be published in indexed journals, in English or Italian.

Two authors independently went through the study selection process using an online electronic systematic review software package (Rayyan QCRI)^37^ to organize and track the process. Any discrepancy was resolved by consensus or by the arbitrary decision of a third review author.

The full search strategy is reported in **Table 1**.

**Table 1.** Applied search terms during systematic

| Databases | Search terms |  |
| --- | --- | --- |
| **MEDLINE**<br>(filters:<br>human;<br>2000-2021) | **#1**<br>**Musculoskeletal disorder** | ((musculoskeletal diseases[MeSH Terms]) OR "musculoskeletal disease*" OR "musculoskeletal disorder*" OR "musculoskeletal dysfunction*" OR "musculoskeletal condition*" OR ("Musculoskeletal Pain"[Mesh]) OR "musculoskeletal pain" OR (neuromuscular diseases[MeSH Terms]) OR "neuromuscular disease*" OR "neuromuscular dysfunction*" OR "neuromuscular disorder*" OR (wounds and injuries[MeSH Terms]) OR injur* OR sprain* OR strain* OR (tendinopathies[MeSH Terms] OR thendinopath*) OR (syndrome[MeSH Terms]) OR syndrome* OR overuse* OR impingement* OR trauma OR "repeated trauma*" OR microtrauma* OR instability OR degeneration OR aspecific OR "non-specific" OR "associated disorder*")<br>AND |
|  | **#2**<br>**Central sensitization** | ((Central Nervous System Sensitization[MeSH Terms]) OR (Somatosensory Disorders[MeSH Terms]) OR "central sensitization" OR "widespread pain" OR "central sensitisation" OR "central hypersensitivity" OR "central nervous system hypersensitivity" OR "central nervous system hyperexcitability" OR "central hyperexcitability" OR "central hyperalgesia" OR "primary hyperalgesia" OR allodynia OR (postsynaptic potential summation[MeSH Terms]) OR "postsynaptic potential summation" OR "quantitative sensory testing" OR "conditioned pain modulation" OR "suprathreshold heat pain response" OR "temporal summation" OR "spatial summation"))<br>AND |
|  | **#3**<br>**Comorbidity** | (Cancer OR tumor OR neoplasm* OR "rheumatoid arthritis" OR fibromyalgia OR "heart failure" OR "ischemic heart disease" OR hypertension OR "type 2 diabetes" OR depression OR anxiety OR "psychological stress" OR "musculoskeletal disease*" OR stroke OR "multiple sclerosis" OR (nervous system disease[MeSH Terms]) OR (musculoskeletal diseases[MeSH Terms]) OR (neuromuscular diseases[MeSH Terms]) OR (depression[MeSH Terms]) OR (anxiety[MeSH Terms]) OR (cardiovascular disease[MeSH Terms]) OR ("Diabetes Mellitus, Type 2"[Mesh]) OR (rheumatic diseases[MeSH Terms]) OR (arthritis[MeSH Terms]) OR (neoplasms[MeSH Terms])) |
| **WEB OF SCIENCE**<br>(filter:<br>2000-2021) | **#1**<br>**Musculoskeletal disorder** | ("musculoskeletal disease*" OR "musculoskeletal disorder*" OR "musculoskeletal dysfunction*" OR "musculoskeletal pain" OR "musculoskeletal condition*" OR "neuromuscular disease*" OR "neuromuscular dysfunction*" OR "neuromuscular disorder*" OR injur* OR sprain* OR strain* OR thendinopath* OR syndrome* OR overuse* OR impingement* OR trauma OR "repeated trauma*" OR microtrauma* OR instability OR degeneration OR aspecific OR "non-specific" OR "associated disorder*")<br>AND |
|  | **#2**<br>**Central sensitization** | ("Central Nervous System Sensitization" OR "Somatosensory Disorders" OR "central sensitization" OR "widespread pain" OR "central sensitisation" OR "central hypersensitivity" OR "central hyperexcitability" OR "central nervous system hypersensitivity" OR "central nervous system hyperexcitability" OR "central hyperalgesia" OR "primary hyperalgesia" OR allodynia OR "postsynaptic potential summation" OR "quantitative sensory testing" OR "conditioned pain modulation" OR "suprathreshold heat pain response" OR "temporal summation" OR "spatial summation")<br>AND |
|  | **#3**<br>**Comorbidity** | (Cancer OR tumor OR neoplasm* OR "rheumatoid arthritis" OR fibromyalgia OR "heart failure" OR "ischemic heart disease" OR hypertension OR "type 2 diabetes" OR depression OR anxiety OR "psychological stress" OR "musculoskeletal disease*" OR stroke OR "multiple sclerosis" OR "nervous system disease*" OR "musculoskeletal disease*" OR "neuromuscular disease*" OR "cardiovascular disease" OR "diabetes Mellitus type 2" OR "rheumatic diseases" OR arthritis) |

### 2.4 Data extraction

Two reviewers independently performed the data extraction process. Any discrepancy was resolved consensually or through the arbitrary decision of a third review author.

The extracted data included: bibliographic information, study design, country, sample size, gender, age, BMI, pathologies, and measurements of signs of CS with their results.

All outcomes were continuous. Therefore, we extracted the mean change from baseline to endpoint (or, alternatively, the mean score at endpoint), the standard deviation (SD) or standard error of these values, and the number of patients included in the analyses.

We asked study authors to supply missing data when necessary.

### 2.5 Methodological assessment

Two independent authors conducted the risk of bias assessment. Any discrepancy was resolved consensually or through the arbitrary decision of a third author.

We assessed the risk of bias (RoB) with the screening tool for cross-sectional studies produced by “The Joanna Briggs Institute Critical Appraisal tools”^38^.

### 2.6 Data analysis

When studies were judged homogenous enough, we performed a random–effect meta-analysis (MA). Otherwise, we reported the results through a qualitative synthesis.

The heterogeneity was assessed using the I^2^ statistics and its interpretation provided by the Cochrane handbook^39^.

The analysis was carried out using RevMan^40^.

## 3. Results

From database investigations, we retrieved 14199 records, 1633 of which were duplicates. We screened the titles and abstracts of the remaining 12566 records. From these, we selected 32 reports for full-text analysis. Ultimately, 14 studies met the inclusion criteria and were included in the SR (see **Fig. 1**).

**Fig. 1.**
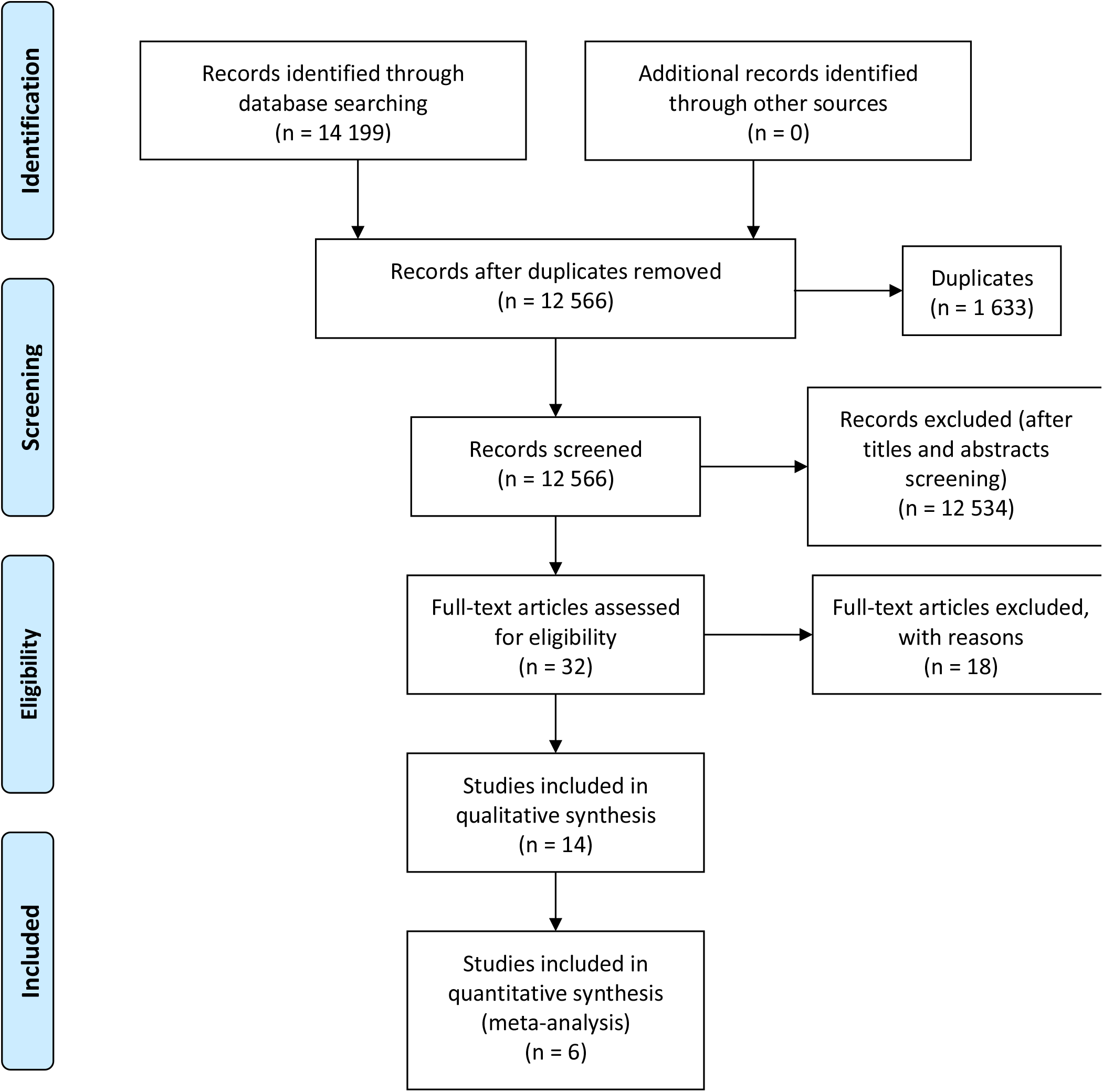
PRISMA Flow Diagram

### 3.1 Characteristics of the included studies

**Table 2** reports the characteristics of the included studies.

**Table 2.** Characteristics of included studies

| Author, year | Study design | Country | Pathologies | Gender | Age | BMI | Outcome | Results |
| --- | --- | --- | --- | --- | --- | --- | --- | --- |
| Aoyagi K et al., 2019 | Cross-sectional study | USA* | 22 CLBP+FMS | 17 F (77.3%) | 43.95±14.00 | 31.34±6.13 | QST: PPT [kg/cm<sup>2</sup>]<br>CPM (PPT-PPT) | PPT thumbnail (CLBP+FMS; CLBP; H): 4.17±2.12; 6.30±3.16; 6.21±1.83; p<0.01.<br>PPT lower back (CLBP+FMS; CLBP; H): 3.14±2.04; 5.80±3.46; 6.82±3.10; p<0.001.<br>CPM thumbnail (CLBP+FMS; CLBP; H): 0.49±0.94; 1.51±1.13; 1.79±0.84; p<0.001.<br>CPM lower back (CLBP+FMS; CLBP; H): 1.05±0.73; 1.72±1.11; 2.42±1.59; p<0.001. |
|  |  |  | 24 CLBP | 15 (62.5%) | 42.38±12.37 | 28.76±6.20 |  |  |
|  |  |  | 22 H | 15 (68.2%) | 41.15±8.83 | 28.35±8.10 |  |  |
|  |  |  | Sample size: 68 | Tot: 47 F (69.1%) | Tot: 42.49 [21-70] | Tot: 29.46 |  |  |
| Bevilaqua-Grossi D et al., 2010 | Cross-sectional study* | Brazil* | 40 TMD+EM | 37 F (92.5%) | 41.30 |  | QST: PPT [g/cm<sup>2</sup>]; h&cPT [°C] | MA |
|  |  |  | 15 EM | 11 F (73.3%) | 45.90 |  |  |  |
|  |  |  | Sample size: 55 | Tot: 48 F (87.3%) | Tot: 42.55 [18-65] |  |  |  |
| Campbell CM et al., 2015 | Case-control study | USA* | 118 KOA+I | 92 F (78.0%) | 59.20±9.10 | 31.20±6.50 | QST: PPT [kPa/cm<sup>2</sup>]; hPT [°C]<br>Dynamic QST: thermal TS, mechanical TS, after-sensation<br>CPM: CPT-PPT, after-sensation | PPT (KOA+I; KOA; I; H): Quadriceps (index knee): 507.60±234.20; 519.20±202.80; 668.10±257.00; 564.70±222.00.<br>Trapezius: 355.90±150.00; 403.40±183.40; 423.10±152.80; 367.30±160.30.<br>TS (KOA+I; KOA; I; H): Forearm-difference score: 15.40±12.50; 14.90±12.80; 13.40±11.60; 12.30±11.10.<br>Knee-difference score: 26.00±20.10; 24.60±16.00; 26.20±19.50; 18.80±20.40.<br>Forearm-difference score: 7.50±11.50; 11.30±12.50; 6.00±8.40; 6.40±6.80.<br>Aftersensation ratings: 19.80±26.90; 6.80±12.40; 11.80±19.70; 10.00±16.20.<br>CMP (KOA+I; KOA; I; H): CPM difference trapezius: 74.40±89.00; 73.80±85.20; 92.40±96.60; 102.70±99.10.<br>Aftersensation ratings: 31.80±21.70; 19.60±18.80; 24.70±17.20; 19.80±18.00. |
|  |  |  | 31 KOA | 16 (51.6%) | 66.55±10.60 | 29.10±4.70 |  |  |
|  |  |  | 30 I | 24 (80.0%) | 58.90±8.60 | 26.80±5.10 |  |  |
|  |  |  | 29 H | 18 (62.1%) | 57.60±7.30 | 26.00±4.80 |  |  |
|  |  |  | Sample size: 208 | Tot: 150 F (72.1%) | Tot: 60.03±9.40 | Tot: 29.53±6.20 |  |  |
| Chaves TC et al., 2016 | Case-control study* | Brazil | 20 TMD+EM | 20 F (100%) | 34.20 (30.41–37.99) | 25.24 | QST: PPT [g/cm<sup>2</sup>]; h&cPT [°C] | MA |
|  |  |  | 20 TMD | 20 F (100%) | 31.65 (25.48–37.82) | 22.05 |  |  |
|  |  |  | 20 EM | 20 F (100%) | 35.05 (29.73–40.37) | 25.77 |  |  |
|  |  |  | 20 H | 20 F (100%) | 26.15 (23.57–28.73) | 22.49 |  |  |
|  |  |  | Sample size: 80 | Tot: 80 F (100%) | Tot: 31.76 [18-65] | Tot: 23.89 |  |  |
| Garrigós-Pedró M et al., 2019 | Cross-sectional study | Spain | 52 TMD+CM | 48 F (92.3%) | 46.20±9.50 | 24.70±4.10 | QST: PPT [kg/cm<sup>2</sup>] | MA |
|  |  |  | 30 H | 24 F (80.0%) | 47.40±10.00 | 24.00±2.70 |  |  |
|  |  |  | Sample size:<br>82 | Tot: 72 F<br>(87.8%) | Tot:<br>46.64<br>[21-65] | Tot:<br>24.44 |  |  |
| Giamberardino MA et al., 2015 | Case-control study* | Italy | 42 EM+FMS | 42 F<br>(100%) | 38.74±<br>5.93 |  | QST: PPT [kgf/cm<sup>2</sup>]; EPT [mA] | Pain thresholds in control areas: the lowest electrical thresholds at all body sites and all tissues and lowest muscle pressure pain thresholds were found in CM+FMS followed by EM+FMS, FMS, CM and EM. The trend for variation among groups was significant for all parameters (p<0.0001). Pain thresholds in tender points: the lowest mean pressure pain thresholds at tender point site were found in CM+FMS followed by EM+FMS, FMS, CM and EM. The trend for variation among groups was significant (p<0.0001). |
|  |  |  | 40 CM+FMS | 40 F<br>(100%) | 37.17±<br>5.73 |  |  |  |
|  |  |  | 41 EM | 41 F<br>(100%) | 38.15±<br>5.37 |  |  |  |
|  |  |  | 40 CM | 40 F<br>(100%) | 36.90±<br>5.61 |  |  |  |
|  |  |  | 40 FMS | 40 F<br>(100%) | 39.92±<br>6.17 |  |  |  |
|  |  |  | Sample size:<br>203 | 203 F<br>(100%) | Tot:<br>38.18<br>[18-65] |  |  |  |
| Janal MN et al., 2016 | Case-control study | USA | 25 TMD+FMS | 25 F<br>(100%) | 43.4±<br>20.40 |  | QST: hPT [°C]<br>Dynamic QST: thermal TS, after-sensation | MA |
|  |  |  | 100 TMD | 100 F<br>(100%) | 36.3±<br>17.30 |  |  |  |
|  |  |  | 48 H | 48 F<br>(100%) | 36.7±<br>14.20 |  |  |  |
|  |  |  | Sample size:<br>173 | Tot: 173 F (100%) | Tot:<br>39.20±<br>14.60<br>[range 19-78] |  |  |  |
| Kaplan CM et al., 2020 | Case-control study* | UK | 27 RA+FMS | 19 F<br>(70.4%) | 52.97±<br>11.07 | 28.22±<br>6.72 | fMRI | In RA+FMS patients: higher levels of peripheral inflammation were associated with increased functional connectivity between the insula and the left midfrontal gyrus (MFG) and the left inferior parietal lobule (IPL); between the left IPL and multiple cortical regions. |
|  |  |  | 27 RA | 22 F<br>(81.5%) | 56.91±<br>11.61 | 26.42±<br>5.10 |  |  |
|  |  |  | Sample size:<br>54 | Tot: 41 F<br>(75.9%) | Tot:<br>54.94 | Tot:<br>27.32 |  |  |
| Munoz-Garcia D et al., 2017 | Cross-sectional study | Spain* | 29 TMD+CNP | 26<br>(89.7%) | 33.21±<br>6.69 |  | QST: PPT [kg/cm<sup>2</sup>] | MA |
|  |  |  | 27 CNP | 17<br>(63.0%) | 27.52±<br>4.82 |  |  |  |
|  |  |  | 30 H | 17<br>(56.7%) | 26.21±<br>4.51 |  |  |  |
|  |  |  | Sample size:<br>86 | Tot: 60 F<br>(69.8%) | Tot:<br>28.98<br>[18-40] |  |  |  |
| Neville SJ et al., 2018 | Cross-sectional study | USA | 3.8% KOA+FMS |  |  |  | QST: PPT [kgf/cm<sup>2</sup>]<br>Dynamic QST: mechanical TS<br>CPM (PPT-PPT) | In females, FM scores correlated with all measures of pressure pain sensitivity (p≤0.021) except thumbnail PPT. These correlations remained significant (p≤0.015) in patients that satisfied survey criteria for FM. No QST outcomes correlated with FM score in males (p>0.05). Other measures (TS and CPM) were not associated to FM score. |
|  |  |  | 96.2% KOA |  |  |  |  |  |
|  |  |  | Sample size:<br>129 | Tot: 68 F<br>(52.7%) | Tot:<br>64.12<br>[≥ 18] |  |  |  |
| Pereira Silva Moreira VM et al., 2017 | Case-control* | Brazil | 15 KOA+KOA | 7 F<br>(46.7%) | 64.00±<br>10.06 |  | QST: PPT [kg/cm<sup>2</sup>] | Significant difference in the mean values of the PPT was found between control and KOA (unilateral and bilateral) patients (p<0.03), while no difference was |
|  |  |  | 15 KOA | 11 F<br>(73.3%) | 59.86±<br>7.61 |  |  |  |
|  |  |  | 10 H | 6 F<br>(60.0%) | 57.80±<br>6.22 |  |  |  |
|  |  |  | Sample size:<br>40 | Tot: 24 F<br>(60.0%) | Tot:<br>60.90<br>[≥ 50] |  |  | found between KOA unilateral<br>and bilateral groups. |
| Peres<br>MFP et<br>al., 2004 | Case-control<br>study* | Brazil* | 12 CM+FMS | 9 F<br>(75.0%) | 42.00±<br>13.18 |  | Cerebrospinal<br>fluid glutamate<br>levels [mmol/l] | CM+FMS patients had<br>significantly higher CSF glutamate<br>levels than patients without<br>fibromyalgia (0.34±0.27 vs<br>0.19±0.06 µmol/l) (p<0.04). Both<br>groups had higher levels than<br>controls p<0.001). |
|  |  |  | 8 CM | 5 F<br>(62.5%) | 42.63±<br>10.69 |  |  |  |
|  |  |  | 20 H |  |  |  |  |  |
|  |  |  | Sample size:<br>40 |  | [range<br>18-64] |  |  |  |
| Sato H et<br>al., 2012 | Case-control | Japan* | 13 TMD+TTH | 13 F<br>(100%) | 28.80±<br>9.80 |  | Dynamic QST:<br>thermal TS,<br>after-sensation | For each stimulus protocol, there<br>were no significant differences<br>between the two groups or<br>between the two stimulus sites<br>with increasing number of stimuli<br>(low-stimulus protocol, F=0.258,<br>p=0.773; high-stimulus protocol,<br>F=0.216, p=0.806). |
|  |  |  | 20 H | 20 F<br>(100%) | 22.95±<br>4.60 |  |  |  |
|  |  |  | Sample size:<br>33 | Tot: 33 F<br>(100%) | Tot:<br>25.25 |  |  |  |
| Smith MT<br>et al.,<br>2009 | Cross-sectional<br>study* | USA | 36 TMD+I |  |  |  | QST: PPT<br>[kPa/cm<sup>2</sup>]; hPT<br>[°C] | MA |
|  |  |  | 17 TMD |  |  |  |  |  |
|  |  |  | Sample size:<br>53 | Tot: 43 F<br>(81.1%) | Tot:<br>33.60±<br>12.40 | Tot:<br>25.40±<br>5.30 |  |  |

\*: data not explicitly expressed in the study, but inferred by the reviewers
AS: after-sensation;
CLBP: chronic low back pain;
CM: chronic migraine;
CNP: chronic neck pain;
CPM: conditioned pain modulation;
CPT: cold pressure test;
CSF: cerebrospinal fluid;
EC: episodic migraine;
EPT: electrical pain threshold;
F: female;
fMRI: functional magnetic resonance;
FMS: fibromyalgia syndrome;
H: healthy;

All the included studies were analytical observational studies: six cross–sectional studies^41–46^ and eight case–control studies^47–54^. One^52^ was published in 2004 and another^46^ in 2009, while all the others^29–40^ have been published in the last decade.

The sample sizes ranged from 33 to 208 patients. All studies included more female than male, with four studies that recruited only women. The total number of subjects assessed was 1304, of which 1056 were women. The mean age of recruited patients was 45.4, ranging from 18 to 78 years old. TMD^42–44,46,47,49,53^, knee OA (KOA)^45,51,54^, CLBP^41^, RA^48^, and migraine^50,52^ are found as primary disorders (MSK-D or migraine) in the samples included in this SR. TTH^53^, migraine^42,43,47^, chronic neck pain (CNP)^44^, insomnia^46^, and FMS^49^ are found as comorbidities in patients with TMD. Insomnia^54^, FMS^45^, and controlateral KOA^51^ are found as comorbidities in patients with KOA. Lastly, in samples of subjects with migraine^50,52^, CLBP^41^, and RA^48^, the included comorbidity is FMS, that was the most frequently comorbidity found . The most present MSK-D was TMD, investigated in seven studies. No studies investigated samples with more than one chronic comorbidity in association with the primary disorder (MSK-D or migraine). As explained above, any type of headache and persistent MSK-D, since they comprise the only necessary feature of chronic pathology, have also been included among the comorbidities. Therefore, two studies include samples with two persistent MSK-D present at the same time (TMD and CNP^44^, bilateral KOA^51^).

Eleven authors^27–30,32–36,38,39^ used quantitative sensory testing (QST), which evaluates the PPT, thermal pain threshold (TPT: cold-cPT and heat-hPT), and electrical pain threshold (EPT), to measure CS. Four studies^45,49,53,54^ used dynamic QST (three thermal TS and two mechanical TS) to assess the CS’s signs. In three studies^41,45,54^ the conditioned pain modulation (CPM) was used. Kaplan et al.^48^ and Peres et al.,^52^ on the other hand, used different methods to measure signs of CS: the first one used functional magnetic resonance imaging (fMRI) while the second used the cerebrospinal fluid (CSF) glutamate levels.

### 3.2 Methodological assessment of the included studies

**Table 3** reports the methodological assessment of the included studies.

**Table 3.**
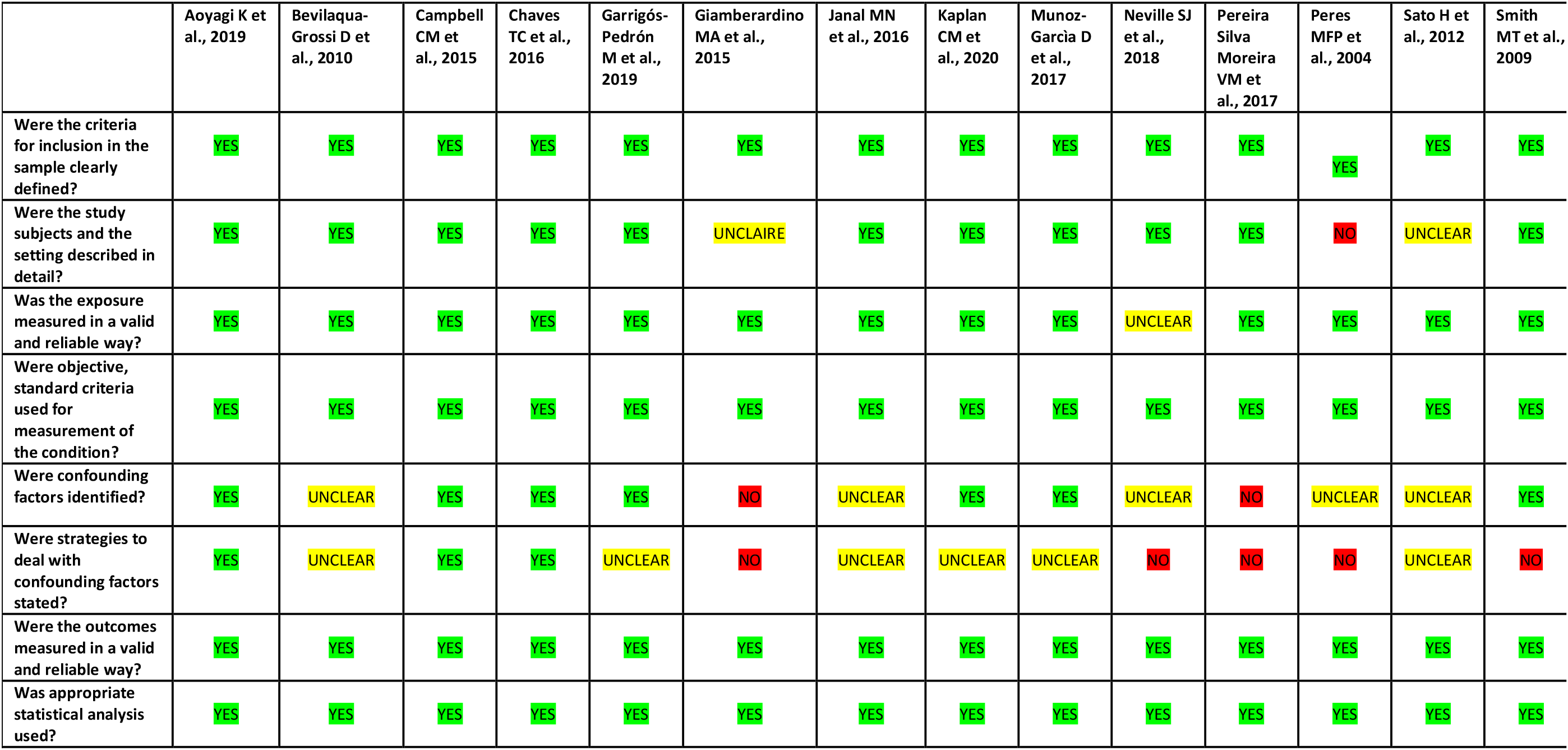
Risk of bias assessment of observational studies

Overall, all the studies resulted in a low or moderate risk of bias.

Inclusion criteria, conditions assessment, outcome measurement, and statistical analysis were evaluated at low risk of bias for all the included studies.

The item “exposure measurement” was judged “unclear” in Neville et al.^45^ Regarding the items on “study subjects” and “setting”, we judged them “not applicable” in Giamberardino et al.,^50^ Sato et al.,^53^ and Peres et al.^52^ Moreover, “confounding factors” and “strategies to deal with confounding factors” were judged inconsistently: in five^42,45,49,52,53^ studies they were not clearly defined, whereas Giamberardino et al.^50^ and Pereira Silva Moreira et al.^51^ omitted them entirely. Finally, “strategies to deal with confounding factors” were not clearly defined by three other authors^43,44,48^ and not reported by Smith et al.^46^

### 3.3 Summary of the results

#### 3.3.1 Temporomandibular disorder

Seven^42–44,46,47,49,53^ studies measured signs of CS in patients with TMD and four different chronic comorbidity: headache (migraine or TTH), insomnia, FMS, and CNP. All studies were similar regarding inclusion criteria. Six^42–44,46,47,49^ studies were homogenous regarding the modality of assessment. Thus, it was possible to performed fifteen different MA aggregating the results of these papers (see **Fig. 2**). Based on the results of the statistical analysis, the estimated Standardized Mean Difference (SMD) in the PPT measurement was decreased in patients with TMD and associated comorbidities versus healthy subjects and was -1.10 [Confidence Interval

**Fig. 2.**
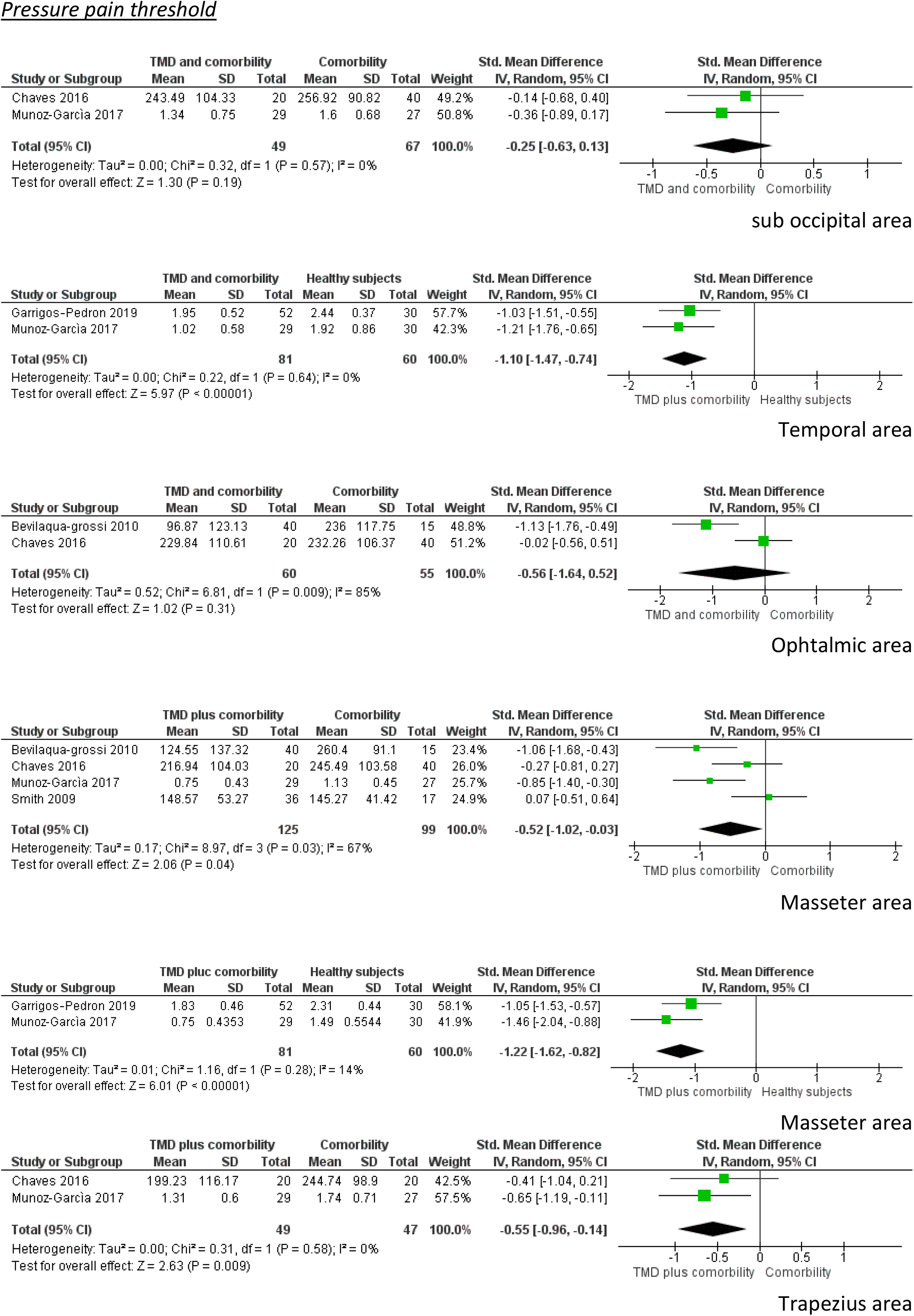
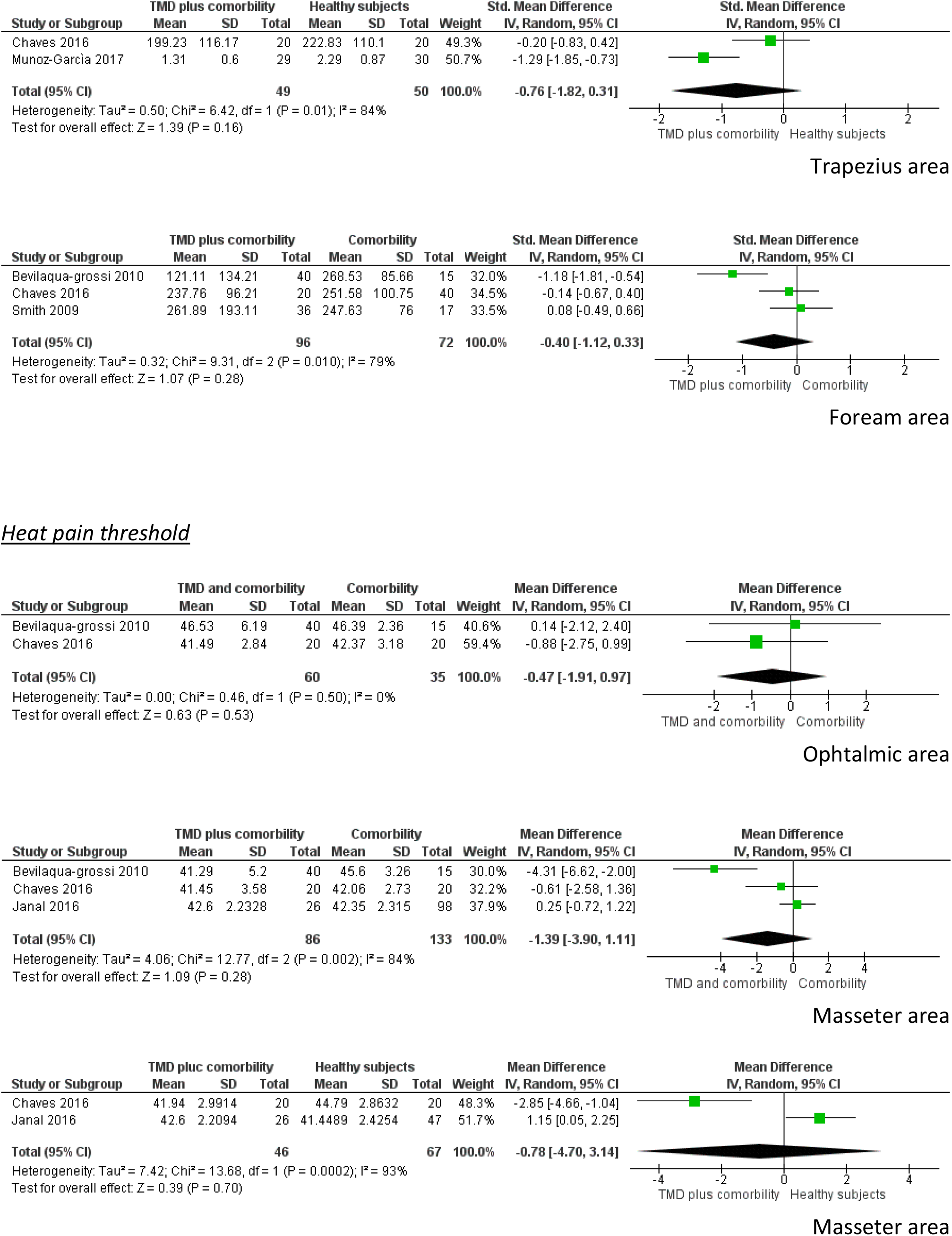
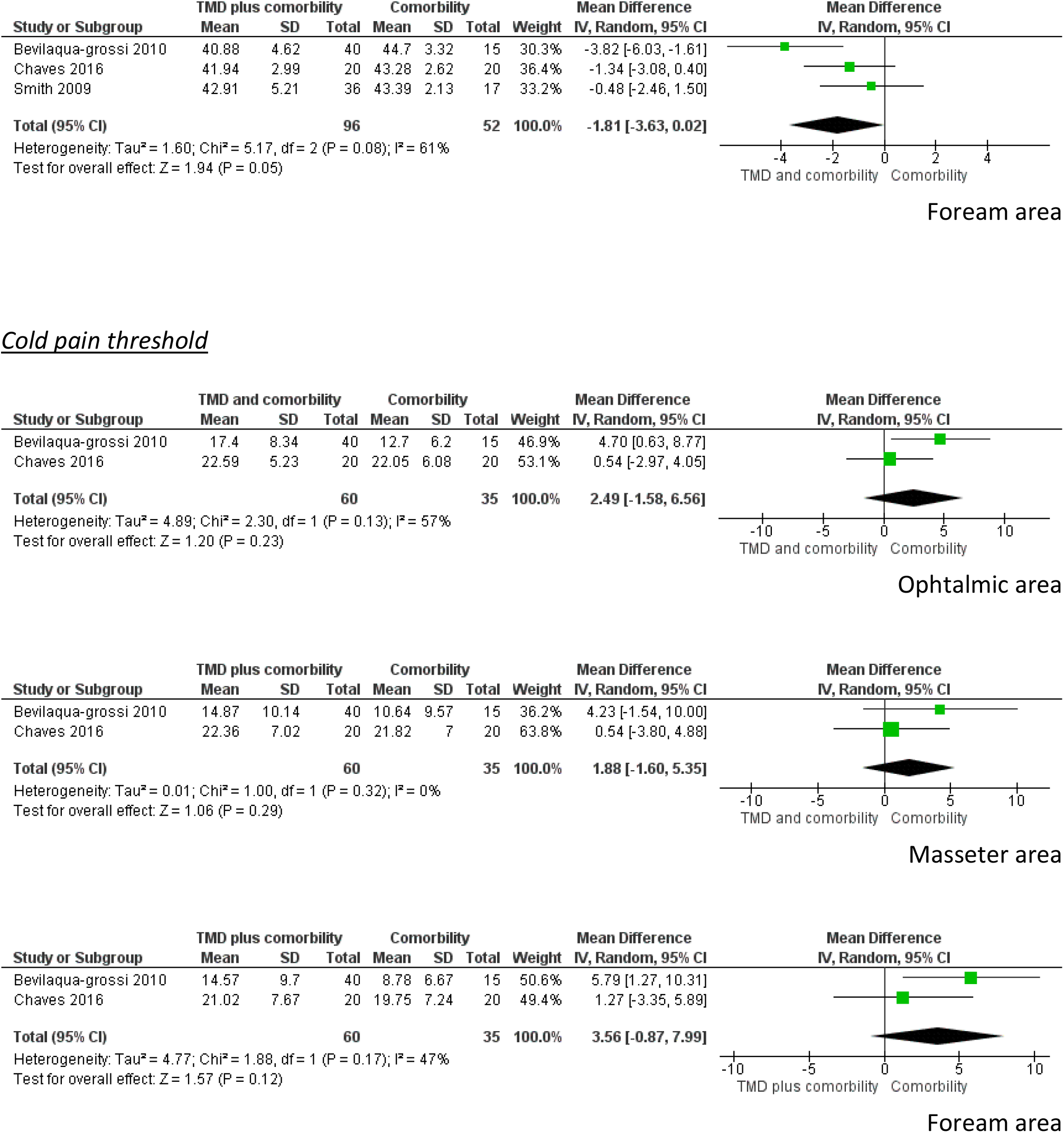
Forest plot of comparison between MSK-D and comorbility patients vs comorbility patients/healthy subjects for presence of signs of CS. SD, standard deviation; IV, inverse variable; CI, confidence intervals

(CI) 95%: -1.47 to -0.74; I^2^: 0%] in the temporal area and -1.22 [CI 95%: -1.62 to -0.82; I^2^: 14%] in the masseter area. These data confirm higher pain sensitivity in the group with TMD and chronic comorbidity. When comparing patients with TMD and associated comorbidities to subjects with only TMD, the SMD in the PPT measure were statistically significantly lower in favor of the first group mentioned in two body regions: estimated data were -0.52 [CI 95%: -1.02 to -0.03; I^2^: 67%] in the masseter area, and -0.55 [CI 95%: -0.96 to -0.14; I^2^: 0%] in trapezius area. Instead, when the latter measure is calculated by comparing patients with TMD and chronic comorbidities and healthy subjects, the CI intersected zero making the lower PPT value not statistically significant in the trapezius area [-0.76 with 95% between -1.82 and 0.31]. When PPT is measured in the suboccipital, ophthalmic and forearm regions and compared between subjects with TMD and patients with TMD and associated comorbidities, the point values are decreased in all regions in the second group mentioned showing more signs of CS, but no estimate is statistically significant.

In TPT measurements, all MA compare patients with TMD and chronic comorbidities with subjects with TMD only. All cPT data were increased in samples with associated comorbidities, possibly indicating more CS, but no point measures were statistically significant. The same trend is repeated in hPT measurements, with point values this time decreased in subjects with associated comorbidities and it had a borderline statistical significance with a p < 0.05 and an estimated SMD of -1.81 [CI 95%: -3.63 to 0.02; I^2^: 61%] only in the remote region of the foream.

We were unable to include one study^53^ in the meta-analysis. Sato et al.^53^ compared TMD and TTH to healthy subjects, and found that temporal summation was not induced in either group when assessed in right cheek and right arm (see **Table 2**).

#### 3.3.2 Knee osteoarthritis

Campbell at al.^54^ reported a significant difference on the CS measure (QST) between KOA patients with insomnia, KOA patients, patients with insomnia and healthy subjects, being the first group the most pain sensitive (p = 0.01): they were significantly more sensitive when compared to healthy controls (p = 0.002) and marginally when compared with patients with insomnia (p = 0.06). In individual laboratory tests, PPT at the index knee appeared different between groups, with the KOA and insomnia group being most sensitive (see **Table 2**).

Moreover, Pereira Silva Moreira at al.^51^ found a statistically significant difference in the mean values of the PPT between healthy subjects compared to KOA patients regardless of unilateral or bilateral presentation of condition, while they found no difference between patients with unilateral KOA and bilateral KOA in the same measures. Patients with KOA had lower PPT in the dermatomes, the myotomes, and the sclerotomes evaluated except that in the sclerotomes of the supraspinous ligaments (see **Table 2**).

Finally, Neville et al.^45^ evaluated patients with KOA and FMS. Their results report that in females, the presence of FMS correlated with all static pain threshold measures (trapezius and tibialis anterior) of pressure pain sensitivity (all p ≤ 0.021) except thumbnail PPT. On the other hand, none of the QST outcomes correlated with the presence of FMS in males (all p > 0.05). CPM was not related to FMS in either sex (see **Table 2**).

#### 3.3.3 Migraine

Peres et al.^52^ reported that migraine patients with FMS had significantly higher CSF glutamate levels than subjects with migraine but without FMS (see **Table 2**). Both these groups had higher levels of CSF glutamate than healthy controls (see **Table 2**).

Giamberardino et al.^50^ compared chronic migraine and FMS, episodic migraine and FMS, FMS, chronic migraine, and episodic migraine patients in the measure of PPT and EPT. They found a trend for lower pain threshold in all sites evaluated among groups for all parameters in both measures (see **Table 2**).

#### 3.3.4 Chronic low back pain

Only Aoyagi et al.^41^ has compared patients with CLBP associated with FMS, with patients with CLBP only and healthy controls. The paper demonstrated decreased measures of PPT and CPM in all sites evaluated (thumbnail and lower back) with statistical significance (see **Table 2**).

#### 3.3.5 Rheumatoid arthritis

Kaplan et al.^48^ demonstrated more signs of CS in a subgroup of subjects with FM and RA, compared to controls with RA only, when evaluated with fMRI (see **Table 2**).

## 4. Discussion

### 4.1 Central sensitization in people with MSK-D and chronic comorbidities

In this SR we tried to estimate whether patients with MSK-D and chronic comorbidities showed more signs of CS, thus demonstrating a nociplastic component in pain and manifest signs and symptoms.

Among the MSK-D, we found only studies with samples with persistent MSK-D and associated comorbidities. Currently, there are no data in the literature on the presence of CS in subjects with a chronic condition who have suffered an acute MSK-D. Therefore, the results of our work are not generalizable to acute MSK-D. The variability in the comorbidities assessed in the included studies is very low, with persistent MSK-D, FMS, insomnia, and migraine as the only comorbidities reported. This does not allow to generalize the results obtained to chronic comorbidities belonging to other systems, such as cardiovascular or metabolic pathologies.

Some papers have already investigated the presence of signs of CS in patients with persistent MSK-D^3–6,15,17^ and in every single disease included among comorbidities.^55–57^ For this reason, the presence of signs of CS appears proven in patients with persistent musculoskeletal symptoms or with complex conditions (i.e., associated comorbidities), confirming the hypothesis initially supported by us and confirmed by the results of this SR. The data resulting from this SR must be interpreted with caution because the two studies considering migraine are of moderate methodological quality; only one study per pathology was found for CLBP and RA. There is great heterogeneity from the point of view of the methodological quality and the characteristics of the included samples in the studies examining subjects with KOA. Finally, only 14 studies were included in the SR, and it was possible to produce MA in only one subgroup of patients.

Previous studies have tried to demonstrate the association of CS in maintaining symptoms and achieving worse outcomes after treatment in MSK-D population. The study by Slade et al.^58^ seems to demonstrate that the presence or absence of signs of CS (PPT) is a consequence of the patient’s painful state, fluctuating in conjunction with the trend of pain levels and does not seem to predict persistent symptom states when initially measured in a cohort of healthy subjects who will then develop temporomandibular pain. The same results appear to be confirmed in a cohort of patients with acute low back pain^59^ not being the main cause of the development of persistent symptoms when considered alone. It could become the cause of persistent pain states when signs of CS are detected in patients with concomitant psychological and psychosocial factors^60,61^ and to an initial state of more intense symptoms.^62^ This statement also finds support after an acute trauma such as whiplash.^63,64^ Clark et al.^65^ detect “sensory hypersensitivity and somatization pre-morbidly, or higher sensory at the acute stage of pain are predictors of altered central pain modulation in some musculoskeletal pain conditions”.

Having evaluated data transversely in this SR, no conclusions can be drawn regarding CS, i.e. whether it is caused by the presence of chronic comorbidities in subjects with MSK-D or whether associated comorbidities simply maintain it after the acute state of the injury in which it may be a physiological protective mechanism. Furthermore, there seems to be an interaction between CS and psychosocial factors, but these are only a small part of all the comorbidities that can involve a patient, and we do not know if these interactions are also common to non-psychological pathologies and so of the health state of the patient. Experimental studies have shown that having suffered a previous injury may be sufficient to establish CS processes,^66^ as well as being exposed to frequent painful stimuli,^67^ and numerous pathologies present pain among the manifest symptoms. Preliminary data seem to be confirmed that identify a causality of the state of health in the maintenance of CS^25^ and, as reported by Neville et al.,^45^ the presence of FMS (and, in general, of chronic comorbidities) may provide a helpful measure of centralized pain processing.

On the other hand, when a subgroup of patients with persistent MSK-D is examined, the presence of signs of CS seems to be present and of marginal importance in giving indications on the course of symptoms, as these will, in any case, only improve to a limited extent.^68,69^ Probably, this reason justifies the slightest difference that emerges from this SR in signs of CS found between samples with two persistent MSK-D present at the same time and subjects with only one MSK-D.^44,51^

### 4.2 Clinical implications

There is more assurance in the data that show that the presence of signs of CS before the implementation of a treatment negatively influences the outcome when measured in terms of pain and disability.^70,71^ Patients with more signs of CS, also show increased and persistent pain after surgery.^72^ O’Leary et al.,^73^ in their SR, confirm that people with signs of CS at baseline, have worse outcomes after treatment (surgical or conservative intervention), although the quality of the evidence is low.

This information is essential in addressing the treatment of these patients, and it is necessary to propose appropriate treatments in order to achieve better results. Due to the presence of signs of CS in subjects with MSK-D and chronic comorbidities, the clinicians must offer treatments to improve psychosocial factors and promote a healthy and active lifestyle, as these are the main factors that contribute to the persistence of symptoms.^74^ The conservative approach^75^ and the physical therapy^76^ were shown to be effective in treating persistent MSK-D, reducing hyperalgesia, and causing brain plastic changes. The non-invasive management should include communication and pain education.^77,78^ As demonstrated by Bricca et al.,^79^ when patients have more than one pathology, the primary way to manage their conditions is the active approach based on physical exercise. Rice et al.,^80^ reported that physical exercise decreases pain sensitivity in healthy controls. This phenomenon is called “exercise-induced hypoalgesia” (EIH). The results are more variable in the chronic pain population (with higher CS signs) and may be impaired in some of this, with pain sensitivity remaining unchanged or increasing in response to exercise.^80–82^

These results highlight how personalized and multimodal the management of MSK-D patients with high CS signs should be.

### 4.3 Research implications

Future research should investigate, with high methodological quality, the presence of signs of CS in patients with acute musculoskeletal injuries and chronic comorbidities. The associated disorders should differ from persistent MSK-D, headache, FMS or sleep disorders. By comparing future results with subjects that present only one acute injury and with data coming from this SR, the correlation between health status and CS could be interpreted more clearly. This would increase the knowledge about the cause of the persistence of symptoms in these patients. There is also a lack of high-quality, long-term prospective studies to clarify the topic. Lastly, it would be useful to investigate how the alterations in central pain modulation worsened by chronic comorbidities may affect the treatment outcome and prognosis in patients with MSK-D.

### 4.4 Limits

This SR has several limits, especially regarding the identification and selection of the studies and the processing of the results.

The inclusion criteria of the studies are stringent and concordant with the clinical question proposed at the beginning of this SR and were established a priori by registering on the PROSPERO database. However, they only take into consideration studies in English or Italian. The papers included have been published from 2000 and were only searched in two databases, however consistent with the researched study design. These decisions may have made the search strategy less sensitive but allowed for studies with only a moderate to low RoB to be selected.

A list of excluded studies was not produced. No validated tools were used to extract the data, but all relevant data relating to the study design, sample and outcomes were established before the start of the SR and blindly extracted. The authors were contacted, if the data relating to the measurement of the SC signs were not present in the studies or were otherwise obtained from the graphs in the papers. Unfortunately, only two authors replied via e-mail, further limiting the chances of MA that have been conducted only in a subgroup of people, aggregating data from no more than four studies for each single MA. The same screening tool used by Joanna Briggs Institute for cross-sectional designs was applied in other works like this one,^83^ as it is the leading study design for this type of investigation and because the data have been processed transversely.

All research steps, selection, and evaluation of the studies, were done by two blind reviewers, with the help of two experts in the field of the main topic of the SR. The exact process was followed in the data processing part, making this report in line with the best indications for SR.

## Conclusions

In conclusion, according to the newest published studies, patients with MSK-D and chronic comorbidities show increased signs of CS and decreased pain threshold compared to healthy controls. When comparing patients with MSK-D and comorbidities with only MSK-D, if the associated pathology is FMS, headache or sleep disorders, the first group seems to show more significant hyperalgesia to painful stimuli. Patients with persistent MSK-D present signs of CS regardless of chronic comorbidities.

Associated comorbidities could be an important factor in maintaining states of CS and contribute to the persistence of musculoskeletal symptoms.

Therefore, contemporary presence of chronic comorbidities can be used as a sign of CS in patients with MSK-D and identifying patients with a nociplastic component in pain and those with worse outcomes after treatment.

## Data Availability

All data produced in the present work are contained in the manuscript

## Abbreviations

MSK-D: musculoskeletal disorders
OA: osteoarthritis
LBP: low back pain
RA: rheumatoid arthritis
CS: central sensitization
IASP: International Association for the Study of Pain
CSI: central sensitization inventory
PPT: the pressure pain thresholds
TS: temporal summation
TMD: temporomandibular disorders
SR: systematic review
PROSPERO: Prospective Register of Systematic Reviews
PRISMA: preferred reporting items for systematic reviews and meta-analyses
TTH: tension type headache
CLBP: chronic low back pain
FMS: fibromyalgia
CSS: central sensitivity syndrome
WoS: web of Science
BMI: body mass index
SD: standard deviation
RoB: risk of bias
MA: meta-analysis
KOA: knee osteoarthritis
CNP: chronic neck pain
QST: quantitative sensory testing
TPT: thermal pain threshold
cPT: cold pain threshold
hPT: heat pain threshold
EPT: electrical pain threshold
CPM: conditioned pain modulation
fMRI: functional magnetic resonance imaging
CSF: cerebrospinal fluid
SMD: Standardized Mean Difference
CI: Confidence Interval
EIH: exercise-induced hypoalgesia.

## Acknowledgements

Not applicable.

## Research funding

Authors state no funding involved.

## Authors’ contributions

MS and DD contributed to the conception and design of the work that led to the manuscript. MS, DD and MC contributed to the drafting the protocol. MS and DD contributed to the materials/analysis tools. MS, DD and MC contributed to the collection, interpretation of data. DF contributed to the analysis of data. MS and DD completed the majority of draft writing. MC and DF provided critical revisions for the manuscript. All authors read and approved the final manuscript. All authors have accepted responsibility for the entire content of this manuscript and approved its submission.

## Availability of data and materials

All data generated or analyzed during this study are included in this published study. Other information of this study are available from the corresponding author on reasonable request.

## Informed consent

Informed consent has been obtained from all individuals included in this study.

## Ethical approval and consent to participate

Not applicable.

